# Childhood educational, social and health factors associated with the timing and recurrence of NEET status into adulthood: a prospective birth cohort study

**DOI:** 10.64898/2026.10.29.26366844

**Authors:** Esme Elsden, Zayn Rajan, Annie Herbert, Alex S. F. Kwong

## Abstract

**Background:** Being not in education, employment or training (NEET) has far-reaching consequences for both health and economic outcomes. Identifying what contributes to, and can predict, becoming NEET is therefore necessary to mitigate negative downstream outcomes. A single measure of whether a young person is NEET can conceal differences in when disengagement occurs, whether it recurs, and what the antecedents are.

**Methods:** We examined childhood educational, social and health characteristics in relation to three dimensions of later NEET: 1) number of occasions of NEET, 2) age at first observed NEET, and 3) single, sustained or recurrent (repeated with interruptions) patterns, using up to 11 repeated assessments between ages 16 and 33 in the Avon Longitudinal Study of Parents and Children (ALSPAC; imputed analyses, N=8,130). Negative binomial and multinomial models compared minimally, sociodemographically and mutually adjusted associations.

**Results:** After mutual adjustment, higher attainment at age 10-11 (Key Stage 2), number of childhood traumas up to age 17, persistent patterns of childhood conduct problems and adolescent depression remained associated with accumulated, sustained and recurrent NEET (e.g. attainment incidence rate ratio: 0.80, 95% CI: 0.75-0.85). NEET antecedents differed by age at first NEET: the widest range of factors predicted onset before 21, only depression predicted onset at 21–24, and onset at 25–33 was dominated by sex, with women twice as likely to experience being NEET. Family-disadvantage associations attenuated after mutual adjustment; complete-case results were consistent.

**Conclusion:** Conditional associations do not establish causal effects, mediation or individual predictive accuracy. However, educational, social and health needs overlap across different patterns of NEET participation, and these findings highlight the need for early educational and family support, integrated mental-health and employment services, and continuing support after re-entry.

**Key messages:** *What is already known on this topic:* NEET (not in education, employment or training) status is associated with childhood disadvantage and mental-health difficulties, but most evidence captures NEET at a single time point, potentially obscuring the relationship between antecedents with timing and recurrence.

*What this study adds:* Using 11 repeated assessments across two decades of a birth cohort born in 1991/2, this study shows that childhood attainment, trauma, conduct problems and adolescent depression are associated with accumulated, sustained and recurrent NEET, and that the association between sex and NEET reverses direction after age 25.

*How this study might affect research, practice or policy:* These findings support multi-agency approaches to NEET prevention that combine educational and mental-health support before school leaving with continuing support after NEET recurrence, rather than relying on single-point interventions.

## Introduction

An estimated 1,012,000 UK 16-24-year-olds were not in education, employment or training (NEET) between January-March 2026^1^. NEET status is not only a labour-market indicator, but also a marker for current and future health and wellbeing. A systematic review and meta- analysis of 24 studies found that young people who are NEET have substantially higher odds of mental ill-health (odds ratio 1.28, 95% confidence interval 1.06 to 1.54), substance use problems (odds ratio 1.43, 95% confidence interval 1.08 to 1.89) and markedly elevated suicidality (odds ratio 2.84, 95% confidence interval 2.04 to 3.95) than their peers^2^, making NEET status a public-health concern.

Framing NEET status as a societal concern is not confined to the mental-health evidence. Reduced educational and occupational engagement in adolescence is itself patterned by the structural inequalities that drive health inequalities across the UK more broadly, so that NEET status can be both a marker and a further driver of unequal life chances across multiple public-health domains, from mental health to health behaviours and long-term physical health^3^. Youth unemployment and disengagement have also been shown to leave a lasting ‘scarring’ effect on mental health and subsequent labour-market outcomes that persists years after a young person re-enters education or employment, suggesting that the consequences of a NEET episode are not necessarily confined to its duration^4^.

NEET status is nevertheless conventionally measured as a single ever/never at one time- point count, but NEET status encompasses experiences that plausibly differ in cause and consequence. An early critique of the NEET indicator, based on interviews with young people and caseworkers, identified three such groups: brief transitional gaps between stages of education or training; young parents who chose to prioritise caring for their children; and young people with multiple compounding risks such as homelessness, offending, or emotional and behavioural difficulties^5^. Collapsing these into a single headline rate obscures this heterogeneity and can push services towards reactive, target-driven responses that leave some of the highest-need young people, such as young parents, with comparatively little support^3^. The same conflation applies at the individual level. A single ever/never measure averages over differences in when these experiences begin and how often they recur. Distinguishing the timing and repetition of NEET, not only its occurrence, may therefore matter for identifying who needs support and when.

The evidence base nevertheless remains dominated by research that captures NEET status at a single wave rather than its timing or recurrence. A global scoping review of 159 studies identified substantial variation in how NEET and its associated characteristics are measured^6^. Where studies have followed young people over time, they link NEET status to socioeconomic resources, adverse childhood experiences, educational development and mental-health difficulties^7, 9–13^. Veldman and colleagues followed a Dutch cohort from age 11 to 26 and found that early internalising problems predicted NEET status that emerged early (by age 19–22) or late (by age 26), but not NEET status that persisted across waves, whereas low parental socioeconomic status and multiple negative life events predicted persistence specifically^7^. A Norwegian life-course study similarly found different long-term outcomes among young people who were NEET early in adulthood, with some going on to fare well and others experiencing persistent exclusion^8^. Together, these studies show that different childhood exposures predict different NEET patterns rather than NEET status as such. Most of the evidence base, however, does not test this: Pitkänen et al., for example, used Finnish register data to show that family socioeconomic resources were more strongly associated with a single register-based assessment of NEET at age 18 than were adverse childhood experiences such as parental psychiatric hospitalisation, whose associations were largely explained by that same socioeconomic disadvantage^9^; a systematic review and meta- analysis of 43 studies found pooled associations with parental worklessness (odds ratio 1.54, 95% confidence interval 1.17 to 1.91) and low parental education (odds ratio 0.81, 95% confidence interval 0.67 to 0.95, for higher versus lower education)^12^, and a separate review of 41 European studies linked attention-deficit/hyperactivity disorder, depression and psychotic disorders with subsequent NEET status^13^. Both drawn largely from single-wave or status-at-one-timepoint designs that cannot distinguish these patterns. Even the studies that do track trajectories rarely extend past the transition to adulthood, rarely isolate age at first NEET occasion as an outcome in its own right and rarely distinguish single from repeated or intermittent spells^6^.

Education, adversity and mental health are related to one another and to family circumstances, so coefficients are not independent causal effects^14^. To our knowledge, existing longitudinal studies of NEET have not examined genetic risk alongside these childhood antecedents. Incorporating a polygenic score for educational attainment offers a further, largely unexamined predictor of NEET trajectories, and can provide evidence of a causal relationship for this particular factor. Likewise, few studies capture the childhood antecedents of NEET as repeated measures across infancy, childhood and adolescence rather than single assessments, even though the persistence and recurrence of these exposures, not only their presence, may matter for later outcomes.

Therefore, we used repeated ALSPAC assessments from ages 16 to 33 to address three questions. First, which childhood educational, social and health characteristics are associated with accumulated NEET occasions, and how do these associations change with adjustment? Second, what are their associations with age at first observed NEET? Third, which characteristics are associated with single, sustained and recurrent (repeated but with interruptions) NEET patterns?

## Methods

### Materials and Methods

Pregnant women resident in Avon, UK with expected dates of delivery between 1st April 1991 and 31st December 1992 were invited to take part in the study^15-18^. 20,248 pregnancies have been identified as being eligible and the initial number of pregnancies enrolled was 14,541. Of the initial pregnancies, there was a total of 14,676 foetuses, resulting in 14,062 live births and 13,988 children who were alive at 1 year of age. When the oldest children were approximately 7 years of age, an attempt was made to bolster the initial sample with eligible cases who had failed to join the study originally, adding a further 906 pregnancies not in the initial sample (913 additional children enrolled). The total sample size for analyses using any data collected after the age of seven is therefore 15,447 pregnancies, resulting in 15,658 foetuses. Of these 14,901 children were alive at 1 year of age. From the age of 30, there were an additional 101 new participants recruited from 101 pregnancies during Phase V enrolment. The total sample size for analyses using any data collected after the age of 30 is therefore 15,548 pregnancies, resulting in 15,690 foetuses. Of these 15,002 children were alive at 1 year of age. The present imputed analyses included 8,130 participants, with 5,270 in the secondary polygenic-score analyses.

ALSPAC study data were collected and managed using REDCap electronic data capture tools hosted at the University of Bristol^19^. REDCap (Research Electronic Data Capture) is a secure, web-based software platform designed to support data capture for research studies. Ethical approval for the study was obtained from the ALSPAC Ethics and Law Committee and the Local Research Ethics Committees. Informed consent for the use of all data collected was obtained from participants following the recommendations of the ALSPAC Ethics and Law Committee at the time. The completion of a questionnaire, either on paper or online, was considered to be written consent from participants to use their data for research purposes. For the majority of tests undertaken during face-to-face visits, verbal consent was obtained from participants (both parents and children as appropriate) prior to the start of any data collection; however, some tests required the completion of a written consent form. Participants can contact the study team at any time to retrospectively withdraw consent for their data to be used. Study participation is voluntary and during all data collection sweeps, information was provided on the intended use of data. Please note that the study website contains details of all the data that is available through a fully searchable data dictionary and variable search tool: http://www.bristol.ac.uk/alspac/researchers/our-data/.

### Measures

The outcome, NEET was self-reported at up to 11 assessments between ages 16 and 33. We examined the number of observed NEET occasions (0-11), age at first observed NEET (16-20, 21-24 or 25-33), and pattern of observed NEET (single occasion, sustained across consecutive assessments, or recurrent across non-consecutive assessments; participants meeting criteria for both sustained and recurrent patterns were classified as recurrent), as per previous research19. Never-observed NEET was the reference category for multinomial models. These outcomes describe observed participation histories rather than exact durations of unemployment or inactivity. Further information on the derivation of the NEET variables is available in the supplement.

Exposures comprised of sex assigned at birth, ethnicity, maternal education and social class, all assessed at birth (gender identity was not assessed in ALSPAC and could not be examined separately), counts of maternal-depression (measured via the Edinburgh Postnatal Depression Scale^21^ on 8 occasions between child ages of 32 weeks’ gestation and 8 years) and childhood financial-problems (assessed on 5 occasions between child ages of 32 weeks gestation and 7 years^22^), childhood physical illness count up to age 16, number of childhood traumas up to age 17^23^, standardised Key Stage 2 attainment (a routine assessment in the UK at age 10-11), and counts of childhood conduct-problems and hyperactivity, assessed using the Strength and Difficulties Questionnaire^24^ on 5 occasions between child ages 6.5 and 16 years, and adolescent-depression symptom occasions, assessed using the Short Mood and Feelings Questionnaire^25^ on 4 occasions between child ages 10 and 16 years. We classified maternal education as A level or above (post-16 qualifications), O level (secondary-school qualification, typically taken at age 16), or below O level. A standardised educational-attainment polygenic score, derived from a large genome- wide association study of educational attainment, was examined separately^26^.

### Statistical analysis

Negative binomial models estimated incidence rate ratios (IRRs) for accumulated NEET occasions. Multinomial logistic models estimated relative risk ratios (RRRs) for first-observed age category and pattern category (sustained or recurrent), each relative to never-observed NEET. These models included the exposure of interest and were adjusted for the number of completed NEET assessments; when the exposure was polygenic risk score for educational attainment, the model additionally included sex, ten ancestry principal components. Although sometimes described as unadjusted, these are minimally adjusted models. Sociodemographic models included sex, ethnicity, maternal education and social class, alongside assessment count. Fully adjusted models mutually included all non-genetic exposures alongside the exposure of interest and assessment count.

The minimally adjusted models were interpreted as overall observed associations conditional on assessment count. The second models assessed associations also conditional on measured sociodemographic background. The final models assessed associations conditional on the broader measured educational, adversity, and health profile. We emphasised the final models when describing overlap between these domains, while retaining all adjustment stages to make attenuation visible. No stage was interpreted as identifying a causal effect, and changes between stages were not treated as a means of quantifying the extent of confounding or mediation. Multinomial relative risk ratios compare the probability ratio of an outcome category to the reference category across exposure values; they are not hazard ratios.

Missing data were addressed using multiple imputation by chained-equations with 50 imputed datasets, and estimates were combined using Rubin’s rules^27^. Only antecedents and covariates were imputed, NEET status was not. To account for multiple testing across antecedents, p-values were corrected within each outcome using the false discovery rate (FDR) method, and associations are described as FDR-corrected unless stated otherwise. Inference emphasised estimates and confidence intervals rather than solely based on statistical significance. Results across categories were described without claiming formal statistical differences between coefficients.

## Results

Of the 8,130 participants, 20.5% (n=1,669) were observed as NEET on at least one occasion, with 3.5% (n=287) having 2 NEET occasions and 2.4% (n=198) having 3 or more. The number of NEET occasions ranged between 0 and 10 (mean=0.31). Of these, 9.8% first became NEET between ages 16–20, 7.6% between 21–24, and 3.9% between 25–33. By pattern, 14.6% experienced a single NEET occasion, 3.9% sustained NEET, and 2.1% recurrent NEET.

### Which childhood factors are associated with accumulated NEET occasions?

Almost all early-life antecedents predicted a higher count of NEET occasions between ages 16 and 33 years in unadjusted models (Figure 1). The largest associations were for adolescent depressive symptoms (IRR 1.43, 95% CI 1.33–1.53), child and adolescent conduct problems (1.36, 1.29–1.43), low maternal education at birth (1.62, 1.40– 1.86) and the number of childhood traumas (1.31, 1.26–1.37), with higher KS2 attainment associated with fewer NEET occasions (0.69, 0.65–0.73). A higher educational attainment polygenic score was also associated with fewer NEET occasions (0.89, 0.84– 0.95). These estimates were similar when adjusted for sociodemographic characteristics. In the fully adjusted model, most family and social antecedents attenuated towards the null, including maternal depression (1.00, 0.96–1.04), maternal social class (0.92, 0.81–1.06) and financial problems (1.04, 1.00–1.08). Four antecedents remained associated following FDR correction, including number of childhood traumas (1.15, 1.10– 1.21), higher KS2 scores (0.80, 0.75–0.85), conduct problems (1.14, 1.07–1.22) and depressive symptoms (1.23, 1.14–1.32). Hyperactivity attenuated completely (1.04, 0.98– 1.09) despite a strong unadjusted association, distinguishing it from conduct problems measured on the same instrument. Imputed estimates are given in Supplementary Table 1, and complete case results in Supplementary Table 2.

**Figure 1.**
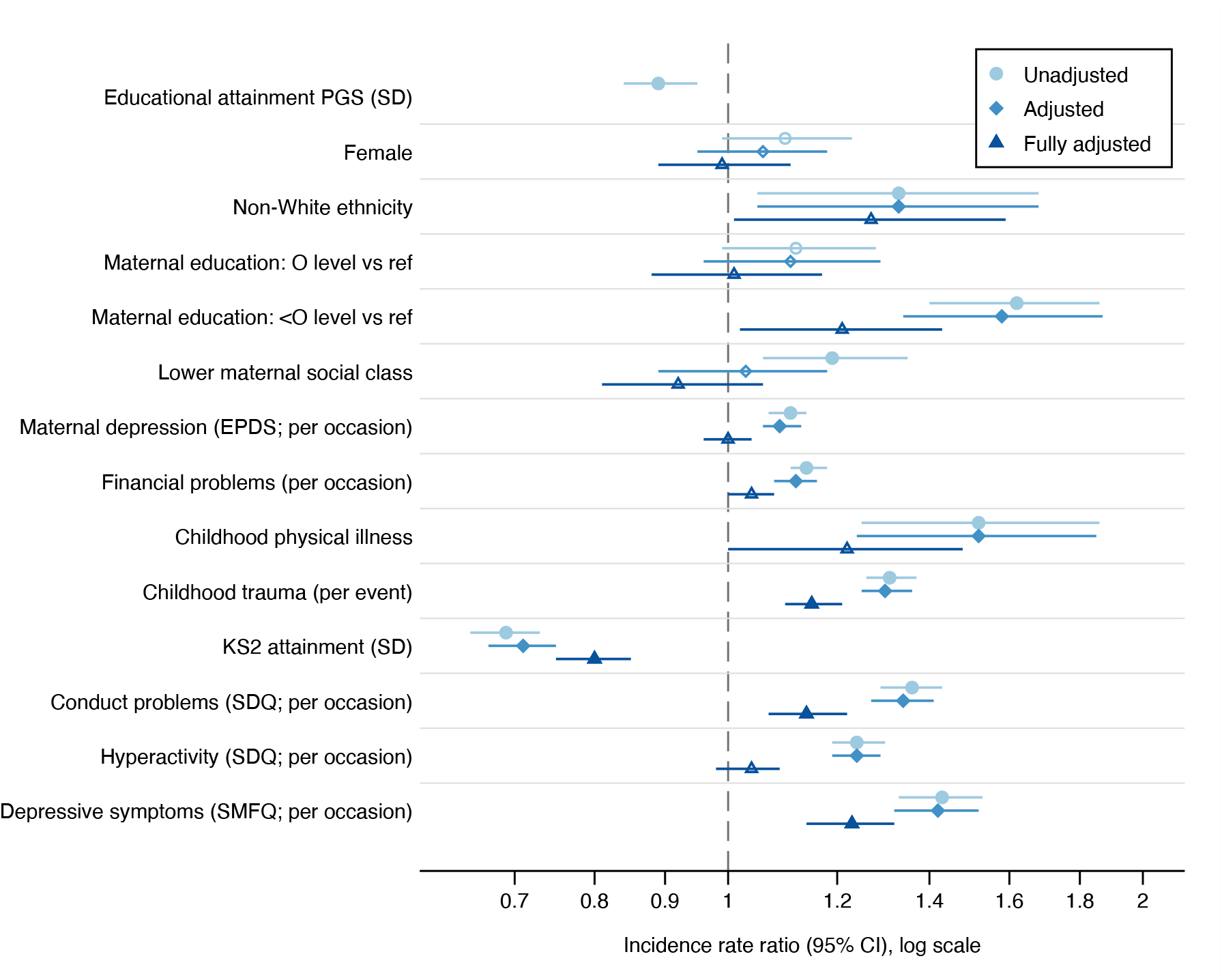
Associations between early-life antecedents and the number of NEET occasions between 16 and 33 years (n=8,130, imputed sample). Incidence rate ratios and 95% confidence intervals from negative binomial regression, shown on a log scale. Light circles, unadjusted; mid-blue diamonds, adjusted for sex, ethnicity and maternal education and social class at birth; dark triangles, fully adjusted for all antecedents simultaneously. All models included the number of NEET questionnaires completed. Filled markers indicate FDR-corrected P < 0.05; hollow markers indicate P ≥ 0.05. The educational attainment polygenic score was available in a subsample (n=5,270) and was adjusted for sex and the first 10 principal components of ancestry.

### How do associations vary by age at first observed NEET?

Antecedents differed markedly by the age at which young people first became NEET (Figure 2). First NEET between 16 and 20 years was predicted by the widest range of exposures: low maternal education (RRR 1.47, 1.15–1.89), financial problems (1.08, 1.02– 1.14), childhood trauma (1.25, 1.16–1.35), KS2 attainment (0.78, 0.71–0.86), conduct problems (1.21, 1.10–1.33) and depressive symptoms (1.23, 1.10–1.38), all still associated following full adjustment. For first NEET between 21 and 24 years, only depressive symptoms were associated (1.26, 1.12–1.42); neither attainment nor childhood adversity predicted entry at this age. First NEET between 25 and 33 years showed a distinct profile again, dominated by sex with women twice as likely to first become NEET in this period (2.01, 1.52–2.65), alongside lower KS2 attainment (0.79, 0.68–0.92), with no surviving associations for trauma, conduct problems or depressive symptoms. The polygenic score was associated with earlier and later onset (0.85 and 0.84) but not with onset between 21 and 24 years (1.05, 0.94–1.16). Imputed estimates are given in Supplementary Table 3, and complete case results in Supplementary Table 4.

**Figure 2.**
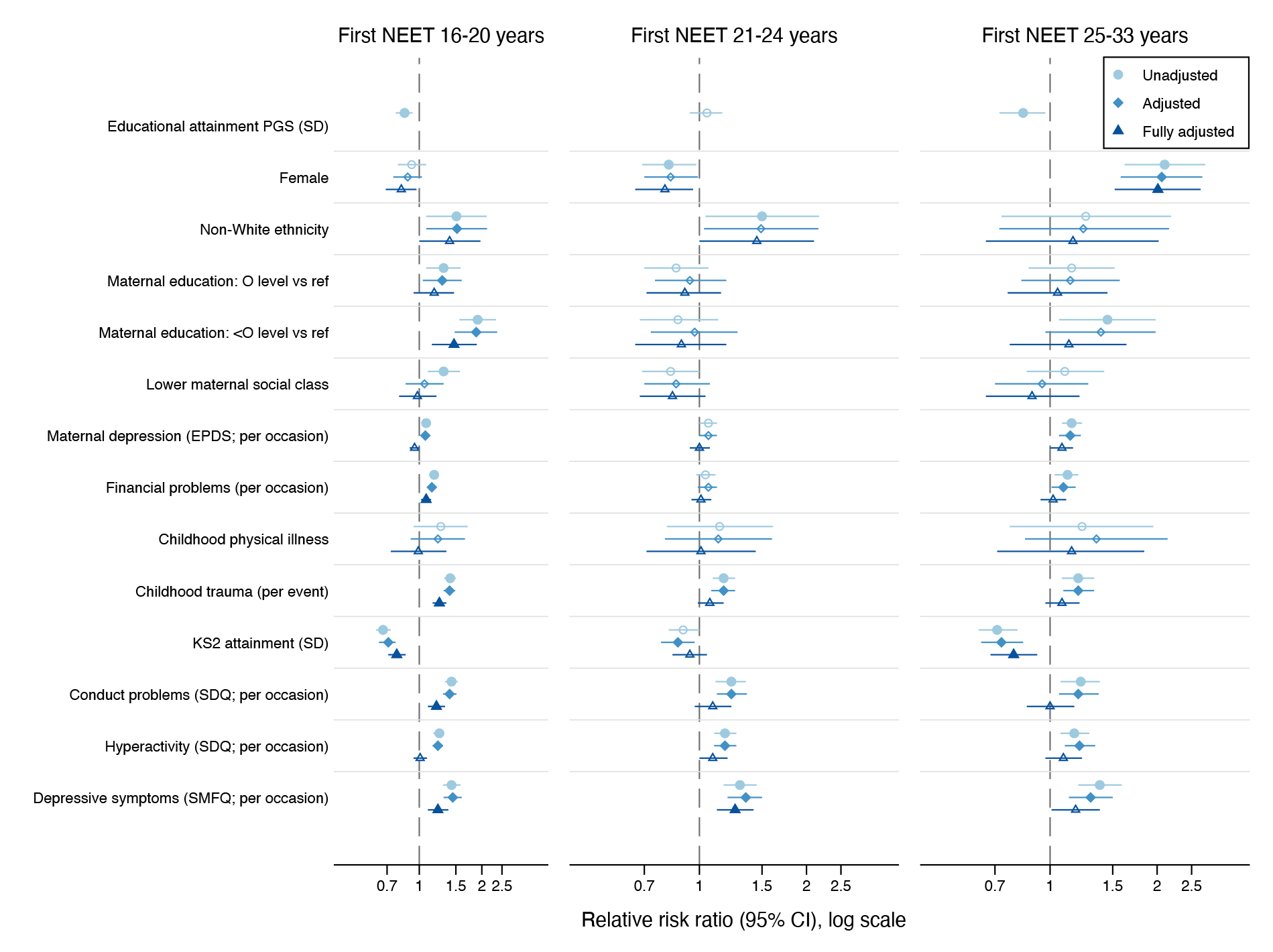
Associations between early-life antecedents and the age of first NEET occasion between 16 and 33 years (n=8,130, imputed sample). Relative risk ratios and 95% confidence intervals from multinomial logistic regression, shown on a log scale. Light circles, unadjusted; mid-blue diamonds, adjusted for sex, ethnicity and maternal education and social class at birth; dark triangles, fully adjusted for all antecedents simultaneously. All models included the number of NEET questionnaires completed. Filled markers indicate FDR-corrected P < 0.05; hollow markers indicate P ≥ 0.05. The educational attainment polygenic score was available in a subsample (n=5,270) and was adjusted for sex and the first 10 principal components of ancestry.

### Which factors are associated with sustained and recurrent NEET?

A similar pattern emerged across NEET types (Figure 3). Single-occasion NEET, the most common form, was weakly predicted overall: only childhood trauma (1.10, 1.03–1.17) and depressive symptoms (1.17, 1.06–1.28) survived adjustment and correction. Sustained NEET showed by far the strongest associations, with higher KS2 attainment the antecedent showing the strongest association (0.65, 0.56–0.75) alongside childhood trauma (1.31, 1.18– 1.45), conduct problems (1.27, 1.12–1.45) and depressive symptoms (1.38, 1.20–1.60). Recurrent NEET was predicted by the same set at more modest magnitude: trauma (1.23, 1.06–1.42), conduct problems (1.28, 1.06–1.54) and depressive symptoms (1.34, 1.11–1.63) — with a large but imprecise association for ethnic minorities (2.08, 1.17–3.72) that fell marginally outside the corrected threshold. The polygenic score was associated with recurrent NEET (0.79, 0.64–0.96) but not with single-occasion NEET (0.95, 0.88– 1.03). Imputed estimates are given in Supplementary Table 5, and complete case results in Supplementary Table 6.

**Figure 3.**
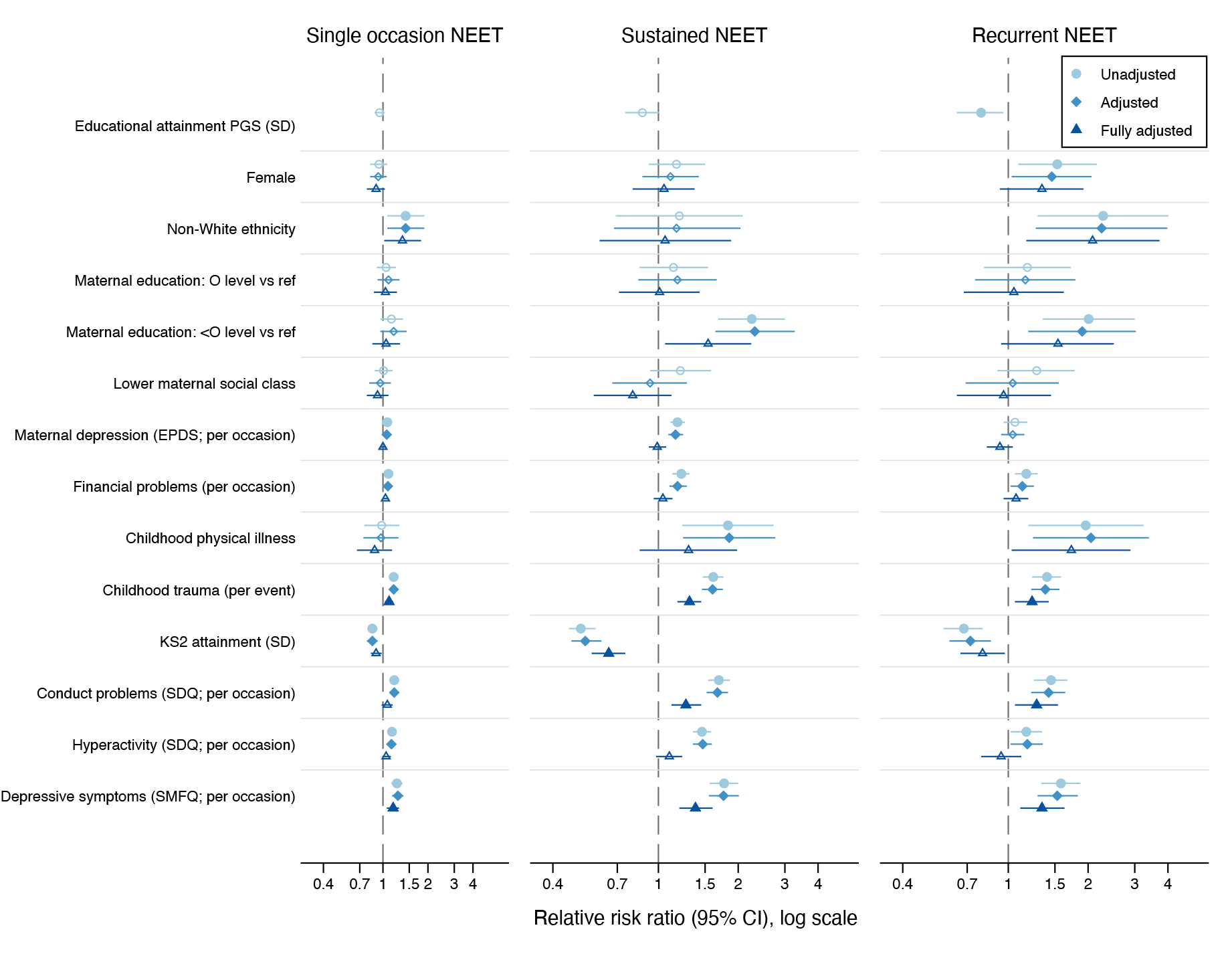
Associations between early-life antecedents and the type of NEET between 16 and 33 years (n=8,130, imputed sample). Relative risk ratios and 95% confidence intervals from multinomial logistic regression, shown on a log scale. Light circles, unadjusted; mid-blue diamonds, adjusted for sex, ethnicity and maternal education and social class at birth; dark triangles, fully adjusted for all antecedents simultaneously. All models included the number of NEET questionnaires completed. Filled markers indicate FDR-corrected P < 0.05; hollow markers indicate P ≥ 0.05. The educational attainment polygenic score was available in a subsample (n=5,270) and was adjusted for sex and the first 10 principal components of ancestry.

### Sensitivity analysis restricted to complete cases

A complete-case analysis restricted to the 4,688 participants with fully observed exposure and covariate data produced fully adjusted estimates closely consistent with the imputed- data results above for all three outcomes (Supplementary Tables S2, S4, S6). Point estimates typically differed by 0.01–0.05 on the ratio scale, confidence intervals overlapped substantially, and no imputed-analysis association reversed direction. The largest difference was for conduct-problem occasions and first observed NEET at ages 25–33 (imputed 1.00, 0.86–1.17; complete case 0.77, 0.58–1.02). Restricting to complete cases therefore did not change which antecedents had associations with NEET.

Across all three sets of models, the same four antecedents recurred: childhood attainment, childhood trauma, conduct problems and adolescent depressive symptoms. Social and economic antecedents such as maternal education, social class, financial hardship and maternal depression were strongly associated with NEET in unadjusted models.

## Discussion

We found that childhood attainment, trauma, conduct problems and adolescent depression were associated with accumulated occasions and with both sustained and recurrent NEET after mutual adjustment. Age at first NEET showed distinct antecedent profiles across age bands, including a sex difference in risk that emerged only from age 25 onwards. The polygenic score predicted recurrence rather than single occasions of NEET. This pattern is consistent with the score indexing more persistent risk rather than any single NEET episode. The polygenic score estimates do not demonstrate a direct genetic effect or validate individual prediction. Together, these findings highlight the need to address early educational and health needs, the circumstances surrounding later disengagement, and support after re- entry. It also reinforces the value of following NEET across adolescence and adulthood, enabling a more nuanced analysis of what antecedents predict accumulation, onset, recurrence and sustained occasions.

The attenuation of family associations should not be interpreted as evidence that NEET is primarily an individual skills or mental-health problem. The first models describe how disadvantage clusters with NEET, whereas the mutually adjusted models condition on characteristics that may themselves reflect earlier social circumstances. A smaller coefficient can result from shared causes, adjustment for intermediate variables, measurement differences or correlated exposures. Without a causal model and appropriate identification assumptions, these changes cannot quantify mediation or isolate direct effects, a distinction sometimes summarised as the Table 2 fallacy^14^. The persistence of associations with low maternal education and financial difficulty further cautions against dismissing material circumstances.

This interpretation complements Gladwell and colleagues’ analysis of the same age range, which found that cognitive ability accumulated across compulsory schooling was the dominant predictor of NEET status^10^. They tested childhood and family factors- socioeconomic status, aspirations, locus of control and risky behaviour- which were associated with NEET indirectly, through their effect on ability formation, rather than directly. They found school engagement and the local youth unemployment rate were the main exceptions, with additional direct associations. Our childhood attainment associations extend across sustained and recurrent patterns. However, attainment is a marker of accumulated learning opportunities and circumstances as well as performance. This leaves out the broader skills acquisition accumulated during school, such as non-cognitive skills^28,29^. Our results do not establish that raising test scores alone would prevent NEET; rather, as Gladwell et al. point to the complex formation of skills needed for future labour transition as a key driver for who becomes NEET or not and indicate a broader systemic issue of what society values and rewards in the labour market^30^.

Our findings contrast with Pitkänen et al., whose Finnish register-based study of 99,137 young people found that parental socioeconomic resources largely accounted for associations between adverse childhood experiences and NEET at age 18^9^. Adjustment for parental education and household income attenuated the odds ratio for parental psychiatric hospitalisation from 1.86 to a non-significant 0.96, and for parental substance-abuse hospitalisation from 2.09 to 1.05, whereas low parental education remained strongly associated (odds ratio 2.94) after full adjustment^9^. Trauma remained associated with accumulated occasions and repeated NEET patterns here. Differences in adversity measures, adjustment sets, national context, institutional culture and follow-up windows may account for this divergence. A Canadian cohort study, born within a similar timeframe to the Finnish cohort above, found support for a specific mediating chain from childhood adversity through parental depression, parent–child conflict and adolescent behavioural difficulties to NEET status at age 24, offering a plausible mechanism for associations we also found even where socioeconomic adjustment attenuates^31^. As the cohorts are not separated by historical era, this contrast is more likely to reflect context-specific differences in social resources and adversity interact over development rather than a universal hierarchy of risk factors.

Adolescent depression was associated with first observed NEET in every age category and with single, sustained and recurrent patterns, and conduct problems also remained associated with repeated NEET. It is highly likely that a large proportion of those in these NEET groups will continue to experience mental health difficulties into adulthood. This argues for mental-health provision embedded alongside educational and employment support rather than offered separately. In England, Mental Health Support Teams (MHSTs) already provide this kind of embedded, early-intervention support within schools and colleges, working alongside education staff rather than through referral to separate clinical services^32^; the 2025 Spending Review committed to extending MHST coverage to all schools and colleges by 2029^33^. Such provision is supported by evidence that NEET young people can remain committed to work while experiencing substantial mental health difficulties^11^. The barrier these associations point to is not necessarily motivation, but access to support that treats mental health and participation together.

Our repeated-pattern findings broadly accord with evidence linking persistent NEET with limited socioeconomic resources and negative life events^7^. Internalising symptom associations concerned early and late NEET, whereas adolescent depression here was associated with both sustained and recurrent classifications. Our study and Veldman et al. differ in observation schedules and definitions; our findings are complementary rather than a direct replication or contradiction. Our findings highlight the need for longer follow-up of NEET in future research in this area, which can reveal participation difficulties that are not captured at the point of leaving school, or even by mid-adulthood.

One policy implication is continued support for coordinated educational, clinical, and employment support, with linked-up placement opportunities to grow skills and confidence. In a randomised trial among young people with first-episode psychosis, individual placement and support improved employment during the intervention^34^. That population differs from this cohort, and the trial cannot establish effectiveness for all young people with depressive symptoms. It nevertheless illustrates why retention, re-entry and wellbeing deserve attention alongside initial job attainment. Our findings motivate testing continued support rather than whether one successful placement resolves recurrent NEET.

The female-sex association with first observed NEET reversed direction after age 25, despite little association with the accumulated count. This finding cautions against treating the school-to-work transition as the only relevant period. Evidence from Denmark demonstrates substantial childbirth-related gender inequalities in employment and earnings^35^, making family formation one plausible explanation. However, these models did not test parenthood, caring responsibilities, or reasons for NEET status. The result cannot be interpreted as evidence that women’s later NEET is caused by motherhood or is necessarily unwanted.

Childcare access, flexible employment and routes back into education or work are therefore hypotheses for further investigation, alongside health-related barriers. Equally, the association between childhood physical illness and recurrent NEET raises questions about accommodations and continuity of support. The aggregated ethnicity result warrants more detailed investigation of unequal opportunities, but cannot identify discrimination or a common experience across diverse ethnic groups. These estimates should inform questions about institutional access rather than individual employability.

A key strength of the study is the repeated assessments and coverage through age 33 that allow several dimensions of NEET to be examined within one cohort. However, the outcomes remain observation based. Consecutive NEET assessments do not establish continuous non-participation between waves, and first observed NEET need not be true onset. Missing waves can alter both the opportunity to observe NEET and its assigned pattern; controlling for assessment count cannot by itself remove informative non-response. The sustained and recurrent groups are descriptive classifications, not estimated transition processes. The age categories for first observed NEET also differ in duration and in the number of underlying assessments, so observed differences between categories may partly reflect unequal opportunity to be classified within them rather than true differences in antecedents.

Some exposure windows also approach or overlap the earliest outcome observations: trauma was measured through age 17, while NEET observation began at 16. Reverse ordering therefore remains possible for early NEET. Attrition, imputation assumptions, residual confounding, and this regional cohort’s historical context constrain inference and generalisability. The many exposure-outcome comparisons warrant caution, particularly where intervals lie near the null. Relative risk ratios also do not describe absolute probabilities, or the likely population benefit of an intervention, rather, the estimated discrepancies in risk between subgroups within the study sample.

In conclusion, this study contributes evidence that educational, social and health needs overlap across repeated patterns of non-participation, while timing reveals additional differences obscured by an overall count. Policies should be evaluated for their ability to sustain participation and wellbeing over time, with support available before school leaving, during later interruptions, and after re-entry.

## Data Availability

The informed consent obtained from ALSPAC (Avon Longitudinal Study of Parents and Children) participants does not allow the data to be made available through any third party maintained public repository. Supporting data are available from ALSPAC on request under the approved proposal number, B5597. Full instructions for applying for data access can be found here: http://www.bristol.ac.uk/alspac/researchers/access/. The ALSPAC study website contains details of all available data (http://www.bristol.ac.uk/alspac/researchers/our-data/).

## Acknowledgements

We are extremely grateful to all the families who took part in this study, the midwives for their help in recruiting them, and the whole ALSPAC team, which includes data collection staff, data and administrative staff, technical managers and the technical staff within the Bristol Bioresource Laboratory, based within the University of Bristol.

## Contributorship statement

E.E. and A.S.F.K. designed the study and acquired funding; A.S.F.K. developed the methodology and analysed the data. E.E. and A.S.F.K. wrote the paper. E.E. and A.S.F.K. are guarantors.

## Ethics approval

This study involves human participants and was approved by the ALSPAC Ethics and Law Committee and Local Research Ethics Committees. Participants gave informed consent to participate in the study before taking part.

