## Supplementary Material for "Childhood educational, social and health factors associated with the timing and recurrence of NEET status into adulthood: a prospective birth cohort study"

**Supplementary Methods**

NEET status was measured over 11 occasions at questionnaires CCS CCT CCU YPA YPB YPC YPE YPG YPJ YPP and the Focus@24 Clinic. We defined NEET status as individuals who reported currently being 1) employed and seeking work, being unable to work through sickness and/or disability, is currently doing voluntary work, or is currently out of work looking after family, at each questionnaire, respectively. We coded individuals at not NEET status if they reported any of the above, alongside being in full or part-time education and/or employment. We derived four measures of NEET from these variables:

First, a count of NEET occasions summed the number of assessments at which a participant met NEET criteria, ranging from 0 to 11. This variable was modelled continuously to estimate the association per additional occasion of NEET.

Second, age at first NEET occasion classified participants according to the age at which they first met NEET criteria: never NEET, first NEET between ages 16 and 20 years, between 21 and 24 years, and between 25 and 33 years. Age bands were defined to correspond broadly to the transition out of compulsory education, early labour market entry, and established adulthood.

Finally, the type of NEET distinguished those never NEET compared to NEET on a single occasion only, those with sustained NEET (NEET at two or more consecutive assessments), or those with recurrent NEET (NEET at two or more non-consecutive assessments, indicating movement in and out of NEET status). Participants meeting criteria for both sustained and recurrent patterns were classified as recurrent.

### Supplementary Table S1

**Supplementary Table 1.** Associations with age at first observed NEET, relative to never-observed NEET. Imputed N=8130. Estimates are RRRs (95% CIs).

| Exposure | Unadjusted | P | Sociodemographic | P | Fully adjusted | P |
|---|---|---|---|---|---|---|
| ***First observed NEET at 16–20 years*** | | | | | | |
| EA polygenic score (per SD) | 0.85 (0.77–0.93) | 0.001 | — | — | — | — |
| Female sex | 0.92 (0.77–0.93) | 0.312 | 0.88 (0.75–1.03) | 0.103 | 0.82 (0.69–0.97) | 0.020 |
| Non-White ethnicity | 1.51 (1.08–2.11) | 0.015 | 1.52 (1.08–2.12) | 0.015 | 1.40 (1.00–1.97) | 0.053 |
| Maternal education: O level | 1.31 (1.08–1.58) | 0.005 | 1.29 (1.04–1.60) | 0.021 | 1.18 (0.94–1.47) | 0.148 |
| Maternal education: below O level | 1.91 (1.56–2.34) | <0.001 | 1.88 (1.48–2.37) | <0.001 | 1.47 (1.15–1.89) | 0.002 |
| Lower maternal social class | 1.31 (1.10–1.57) | 0.002 | 1.06 (0.86–1.31) | 0.574 | 0.98 (0.80–1.21) | 0.873 |
| Maternal-depression occasions | 1.08 (1.04–1.14) | <0.001 | 1.07 (1.02–1.12) | 0.005 | 0.95 (0.90–1.00) | 0.071 |
| Childhood financial-problem occasions | 1.18 (1.12–1.24) | <0.001 | 1.15 (1.10–1.21) | <0.001 | 1.08 (1.02–1.14) | 0.005 |
| Childhood physical illness | 1.27 (0.94–1.71) | 0.113 | 1.23 (0.91–1.66) | 0.172 | 0.99 (0.73–1.35) | 0.965 |
| Childhood traumas | 1.41 (1.32–1.50) | <0.001 | 1.40 (1.31–1.49) | <0.001 | 1.25 (1.16–1.35) | <0.001 |
| KS2 attainment (per SD) | 0.67 (0.62–0.73) | <0.001 | 0.71 (0.64–0.77) | <0.001 | 0.78 (0.71–0.86) | <0.001 |
| Childhood conduct-problem occasions (SDQ-C) | 1.43 (1.33–1.53) | <0.001 | 1.40 (1.30–1.51) | <0.001 | 1.21 (1.10–1.33) | <0.001 |
| Childhood hyperactivity-problem occasions (SDQ-H) | 1.25 (1.17–1.32) | <0.001 | 1.23 (1.16–1.30) | <0.001 | 1.01 (0.94–1.09) | 0.771 |
| Adolescent-depression occasions (SMFQ) | 1.43 (1.30–1.58) | <0.001 | 1.45 (1.31–1.60) | <0.001 | 1.23 (1.10–1.38) | <0.001 |
| ***First observed NEET at 21–24 years*** | | | | | | |
| EA polygenic score (per SD) | 1.05 (0.94–1.16) | 0.387 | — | — | — | — |
| Female sex | 0.82 (0.69–0.98) | 0.025 | 0.83 (0.70–0.99) | 0.035 | 0.80 (0.66–0.96) | 0.015 |
| Non-White ethnicity | 1.50 (1.04–2.17) | 0.031 | 1.49 (1.03–2.16) | 0.035 | 1.45 (1.00–2.10) | 0.053 |
| Maternal education: O level | 0.86 (0.70–1.06) | 0.148 | 0.94 (0.75–1.19) | 0.626 | 0.91 (0.71–1.15) | 0.420 |
| Maternal education: below O level | 0.87 (0.68–1.13) | 0.295 | 0.97 (0.73–1.28) | 0.823 | 0.89 (0.66–1.19) | 0.432 |
| Lower maternal social class | 0.83 (0.69–1.00) | 0.045 | 0.86 (0.70–1.07) | 0.179 | 0.84 (0.68–1.04) | 0.118 |
| Maternal-depression occasions | 1.06 (1.00–1.12) | 0.048 | 1.06 (1.00–1.12) | 0.040 | 1.00 (0.94–1.07) | 0.948 |
| Childhood financial-problem occasions | 1.04 (0.98–1.11) | 0.162 | 1.06 (0.99–1.12) | 0.080 | 1.01 (0.95–1.08) | 0.719 |
| Childhood physical illness | 1.14 (0.81–1.61) | 0.449 | 1.13 (0.80–1.60) | 0.496 | 1.01 (0.71–1.44) | 0.947 |
| Childhood traumas | 1.17 (1.09–1.26) | <0.001 | 1.17 (1.08–1.26) | <0.001 | 1.07 (0.99–1.17) | 0.096 |
| KS2 attainment (per SD) | 0.90 (0.82–1.00) | 0.054 | 0.87 (0.78–0.97) | 0.010 | 0.94 (0.84–1.05) | 0.304 |
| Childhood conduct-problem occasions (SDQ-C) | 1.23 (1.11–1.35) | <0.001 | 1.23 (1.12–1.36) | <0.001 | 1.09 (0.97–1.23) | 0.157 |
| Childhood hyperactivity-problem occasions (SDQ-H) | 1.18 (1.10–1.27) | <0.001 | 1.18 (1.10–1.27) | <0.001 | 1.09 (1.00–1.20) | 0.059 |
| Adolescent-depression occasions (SMFQ) | 1.30 (1.17–1.45) | <0.001 | 1.35 (1.20–1.50) | <0.001 | 1.26 (1.12–1.42) | <0.001 |
| ***First observed NEET at 25–33 years*** | | | | | | |
| EA polygenic score (per SD) | 0.84 (0.72–0.97) | 0.022 | — | — | — | — |
| Female sex | 2.10 (1.62–2.73) | <0.001 | 2.06 (1.58–2.68) | <0.001 | 2.01 (1.52–2.65) | <0.001 |
| Non-White ethnicity | 1.26 (0.73–2.19) | 0.403 | 1.24 (0.72–2.16) | 0.438 | 1.16 (0.66–2.02) | 0.604 |
| Maternal education: O level | 1.15 (0.87–1.52) | 0.341 | 1.14 (0.83–1.57) | 0.420 | 1.05 (0.76–1.45) | 0.763 |
| Maternal education: below O level | 1.45 (1.06–1.98) | 0.019 | 1.39 (0.97–1.98) | 0.073 | 1.13 (0.77–1.64) | 0.534 |
| Lower maternal social class | 1.10 (0.86–1.42) | 0.451 | 0.95 (0.70–1.28) | 0.727 | 0.89 (0.66–1.21) | 0.462 |
| Maternal-depression occasions | 1.15 (1.08–1.23) | <0.001 | 1.14 (1.06–1.22) | <0.001 | 1.08 (1.00–1.16) | 0.046 |
| Childhood financial-problem occasions | 1.12 (1.03–1.20) | 0.004 | 1.09 (1.01–1.18) | 0.025 | 1.02 (0.94–1.11) | 0.654 |
| Childhood physical illness | 1.23 (0.77–1.95) | 0.383 | 1.35 (0.85–2.14) | 0.206 | 1.15 (0.71–1.84) | 0.571 |
| Childhood traumas | 1.20 (1.08–1.33) | <0.001 | 1.20 (1.09–1.33) | <0.001 | 1.08 (0.97–1.21) | 0.165 |
| KS2 attainment (per SD) | 0.71 (0.63–0.81) | <0.001 | 0.73 (0.64–0.84) | <0.001 | 0.79 (0.68–0.92) | 0.002 |
| Childhood conduct-problem occasions (SDQ-C) | 1.22 (1.07–1.38) | 0.002 | 1.20 (1.06–1.37) | 0.005 | 1.00 (0.86–1.17) | 0.970 |
| Childhood hyperactivity-problem occasions (SDQ-H) | 1.17 (1.07–1.29) | <0.001 | 1.21 (1.10–1.34) | <0.001 | 1.09 (0.97–1.23) | 0.166 |
| Adolescent-depression occasions (SMFQ) | 1.38 (1.20–1.59) | <0.001 | 1.30 (1.13–1.50) | <0.001 | 1.18 (1.01–1.38) | 0.039 |

First models adjust for completed assessments; the genetic model also adjusts for sex and ten ancestry principal components. Sociodemographic models add sex, ethnicity, maternal education and social class; fully adjusted models mutually include non-genetic exposures. Maternal education reference: A level or above. Count exposures are per additional occasion, trauma or SD; dashes indicate estimates not available. IRR, incidence rate ratio; RRR, relative risk ratio; CI, confidence interval; EA, educational attainment; KS2, Key Stage 2; SD, standard deviation; SDQ-C, Strengths and Difficulties Questionnaire conduct-problems subscale; SDQ-H, Strengths and Difficulties Questionnaire hyperactivity subscale; SMFQ, Short Mood and Feelings Questionnaire.

### Supplementary Table S2

**Supplementary Table 2:** Associations with accumulated NEET occasions, complete-case analysis (n=4,688; polygenic-score sub-sample n=5,270). Estimates are IRRs (95% CIs).

| Exposure | Unadjusted | P | Sociodemographic | P | Fully adjusted | P |
|---|---|---|---|---|---|---|
| EA polygenic score (per SD) | 0.89 (0.84–0.95) | 0.001 | — | — | — | — |
| Female sex | 1.09 (0.95–1.26) | 0.226 | 1.08 (0.93–1.24) | 0.322 | 1.03 (0.89–1.20) | 0.653 |
| Non-White ethnicity | 1.27 (0.89–1.81) | 0.192 | 1.29 (0.90–1.84) | 0.159 | 1.28 (0.91–1.82) | 0.160 |
| Maternal education: O level | 1.11 (0.95–1.30) | 0.186 | 1.11 (0.93–1.33) | 0.232 | 1.04 (0.88–1.24) | 0.649 |
| Maternal education: below O level | 1.28 (1.05–1.55) | 0.014 | 1.27 (1.02–1.58) | 0.030 | 1.08 (0.87–1.35) | 0.477 |
| Lower maternal social class | 1.09 (0.95–1.25) | 0.233 | 1.00 (0.84–1.18) | 0.966 | 0.93 (0.79–1.09) | 0.383 |
| Maternal-depression occasions | 1.08 (1.04–1.14) | <0.001 | 1.08 (1.03–1.13) | 0.001 | 1.01 (0.96–1.06) | 0.697 |
| Childhood financial-problem occasions | 1.10 (1.05–1.16) | <0.001 | 1.09 (1.04–1.15) | <0.001 | 1.02 (0.97–1.08) | 0.409 |
| Childhood physical illness | 1.41 (1.09–1.81) | 0.008 | 1.43 (1.11–1.84) | 0.005 | 1.19 (0.93–1.53) | 0.163 |
| Childhood traumas | 1.29 (1.22–1.36) | <0.001 | 1.29 (1.21–1.36) | <0.001 | 1.16 (1.09–1.23) | <0.001 |
| KS2 attainment (per SD) | 0.77 (0.71–0.83) | <0.001 | 0.77 (0.71–0.84) | <0.001 | 0.85 (0.78–0.92) | <0.001 |
| Childhood conduct-problem occasions (SDQ-C) | 1.35 (1.24–1.46) | <0.001 | 1.34 (1.24–1.46) | <0.001 | 1.15 (1.05–1.26) | 0.003 |
| Childhood hyperactivity-problem occasions (SDQ-H) | 1.22 (1.15–1.30) | <0.001 | 1.23 (1.16–1.31) | <0.001 | 1.05 (0.98–1.13) | 0.188 |
| Adolescent-depression occasions (SMFQ) | 1.42 (1.29–1.55) | <0.001 | 1.41 (1.28–1.55) | <0.001 | 1.23 (1.12–1.36) | <0.001 |

First models adjust for completed assessments; the genetic model also adjusts for sex and ten ancestry principal components. Sociodemographic models add sex, ethnicity, maternal education and social class; fully adjusted models mutually include non-genetic exposures. Maternal education reference: A level or above. Count exposures are per additional occasion, trauma or SD; dashes indicate estimates not available. IRR, incidence rate ratio; RRR, relative risk ratio; CI, confidence interval; EA, educational attainment; KS2, Key Stage 2; SD, standard deviation; SDQ-C, Strengths and Difficulties Questionnaire conduct-problems subscale; SDQ-H, Strengths and Difficulties Questionnaire hyperactivity subscale; SMFQ, Short Mood and Feelings Questionnaire.

### Supplementary Table S3

**Supplementary Table 3.** Associations with age at first observed NEET, relative to never-observed NEET. Imputed N=8130. Estimates are RRRs (95% CIs).

| Exposure | Unadjusted | P | Sociodemographic | P | Fully adjusted | P |
|---|---|---|---|---|---|---|
| ***First observed NEET at 16–20 years*** | | | | | | |
| EA polygenic score (per SD) | 0.85 (0.77–0.93) | 0.001 | — | — | — | — |
| Female sex | 0.92 (0.77–0.93) | 0.312 | 0.88 (0.75–1.03) | 0.103 | 0.82 (0.69–0.97) | 0.020 |
| Non-White ethnicity | 1.51 (1.08–2.11) | 0.015 | 1.52 (1.08–2.12) | 0.015 | 1.40 (1.00–1.97) | 0.053 |
| Maternal education: O level | 1.31 (1.08–1.58) | 0.005 | 1.29 (1.04–1.60) | 0.021 | 1.18 (0.94–1.47) | 0.148 |
| Maternal education: below O level | 1.91 (1.56–2.34) | <0.001 | 1.88 (1.48–2.37) | <0.001 | 1.47 (1.15–1.89) | 0.002 |
| Lower maternal social class | 1.31 (1.10–1.57) | 0.002 | 1.06 (0.86–1.31) | 0.574 | 0.98 (0.80–1.21) | 0.873 |
| Maternal-depression occasions | 1.08 (1.04–1.14) | <0.001 | 1.07 (1.02–1.12) | 0.005 | 0.95 (0.90–1.00) | 0.071 |
| Childhood financial-problem occasions | 1.18 (1.12–1.24) | <0.001 | 1.15 (1.10–1.21) | <0.001 | 1.08 (1.02–1.14) | 0.005 |
| Childhood physical illness | 1.27 (0.94–1.71) | 0.113 | 1.23 (0.91–1.66) | 0.172 | 0.99 (0.73–1.35) | 0.965 |
| Childhood traumas | 1.41 (1.32–1.50) | <0.001 | 1.40 (1.31–1.49) | <0.001 | 1.25 (1.16–1.35) | <0.001 |
| KS2 attainment (per SD) | 0.67 (0.62–0.73) | <0.001 | 0.71 (0.64–0.77) | <0.001 | 0.78 (0.71–0.86) | <0.001 |
| Childhood conduct-problem occasions (SDQ-C) | 1.43 (1.33–1.53) | <0.001 | 1.40 (1.30–1.51) | <0.001 | 1.21 (1.10–1.33) | <0.001 |
| Childhood hyperactivity-problem occasions (SDQ-H) | 1.25 (1.17–1.32) | <0.001 | 1.23 (1.16–1.30) | <0.001 | 1.01 (0.94–1.09) | 0.771 |
| Adolescent-depression occasions (SMFQ) | 1.43 (1.30–1.58) | <0.001 | 1.45 (1.31–1.60) | <0.001 | 1.23 (1.10–1.38) | <0.001 |
| ***First observed NEET at 21–24 years*** | | | | | | |
| EA polygenic score (per SD) | 1.05 (0.94–1.16) | 0.387 | — | — | — | — |
| Female sex | 0.82 (0.69–0.98) | 0.025 | 0.83 (0.70–0.99) | 0.035 | 0.80 (0.66–0.96) | 0.015 |
| Non-White ethnicity | 1.50 (1.04–2.17) | 0.031 | 1.49 (1.03–2.16) | 0.035 | 1.45 (1.00–2.10) | 0.053 |
| Maternal education: O level | 0.86 (0.70–1.06) | 0.148 | 0.94 (0.75–1.19) | 0.626 | 0.91 (0.71–1.15) | 0.420 |
| Maternal education: below O level | 0.87 (0.68–1.13) | 0.295 | 0.97 (0.73–1.28) | 0.823 | 0.89 (0.66–1.19) | 0.432 |
| Lower maternal social class | 0.83 (0.69–1.00) | 0.045 | 0.86 (0.70–1.07) | 0.179 | 0.84 (0.68–1.04) | 0.118 |
| Maternal-depression occasions | 1.06 (1.00–1.12) | 0.048 | 1.06 (1.00–1.12) | 0.040 | 1.00 (0.94–1.07) | 0.948 |
| Childhood financial-problem occasions | 1.04 (0.98–1.11) | 0.162 | 1.06 (0.99–1.12) | 0.080 | 1.01 (0.95–1.08) | 0.719 |
| Childhood physical illness | 1.14 (0.81–1.61) | 0.449 | 1.13 (0.80–1.60) | 0.496 | 1.01 (0.71–1.44) | 0.947 |
| Childhood traumas | 1.17 (1.09–1.26) | <0.001 | 1.17 (1.08–1.26) | <0.001 | 1.07 (0.99–1.17) | 0.096 |
| KS2 attainment (per SD) | 0.90 (0.82–1.00) | 0.054 | 0.87 (0.78–0.97) | 0.010 | 0.94 (0.84–1.05) | 0.304 |
| Childhood conduct-problem occasions (SDQ-C) | 1.23 (1.11–1.35) | <0.001 | 1.23 (1.12–1.36) | <0.001 | 1.09 (0.97–1.23) | 0.157 |
| Childhood hyperactivity-problem occasions (SDQ-H) | 1.18 (1.10–1.27) | <0.001 | 1.18 (1.10–1.27) | <0.001 | 1.09 (1.00–1.20) | 0.059 |
| Adolescent-depression occasions (SMFQ) | 1.30 (1.17–1.45) | <0.001 | 1.35 (1.20–1.50) | <0.001 | 1.26 (1.12–1.42) | <0.001 |
| ***First observed NEET at 25–33 years*** | | | | | | |
| EA polygenic score (per SD) | 0.84 (0.72–0.97) | 0.022 | — | — | — | — |
| Female sex | 2.10 (1.62–2.73) | <0.001 | 2.06 (1.58–2.68) | <0.001 | 2.01 (1.52–2.65) | <0.001 |
| Non-White ethnicity | 1.26 (0.73–2.19) | 0.403 | 1.24 (0.72–2.16) | 0.438 | 1.16 (0.66–2.02) | 0.604 |
| Maternal education: O level | 1.15 (0.87–1.52) | 0.341 | 1.14 (0.83–1.57) | 0.420 | 1.05 (0.76–1.45) | 0.763 |
| Maternal education: below O level | 1.45 (1.06–1.98) | 0.019 | 1.39 (0.97–1.98) | 0.073 | 1.13 (0.77–1.64) | 0.534 |
| Lower maternal social class | 1.10 (0.86–1.42) | 0.451 | 0.95 (0.70–1.28) | 0.727 | 0.89 (0.66–1.21) | 0.462 |
| Maternal-depression occasions | 1.15 (1.08–1.23) | <0.001 | 1.14 (1.06–1.22) | <0.001 | 1.08 (1.00–1.16) | 0.046 |
| Childhood financial-problem occasions | 1.12 (1.03–1.20) | 0.004 | 1.09 (1.01–1.18) | 0.025 | 1.02 (0.94–1.11) | 0.654 |
| Childhood physical illness | 1.23 (0.77–1.95) | 0.383 | 1.35 (0.85–2.14) | 0.206 | 1.15 (0.71–1.84) | 0.571 |
| Childhood traumas | 1.20 (1.08–1.33) | <0.001 | 1.20 (1.09–1.33) | <0.001 | 1.08 (0.97–1.21) | 0.165 |
| KS2 attainment (per SD) | 0.71 (0.63–0.81) | <0.001 | 0.73 (0.64–0.84) | <0.001 | 0.79 (0.68–0.92) | 0.002 |
| Childhood conduct-problem occasions (SDQ-C) | 1.22 (1.07–1.38) | 0.002 | 1.20 (1.06–1.37) | 0.005 | 1.00 (0.86–1.17) | 0.970 |
| Childhood hyperactivity-problem occasions (SDQ-H) | 1.17 (1.07–1.29) | <0.001 | 1.21 (1.10–1.34) | <0.001 | 1.09 (0.97–1.23) | 0.166 |
| Adolescent-depression occasions (SMFQ) | 1.38 (1.20–1.59) | <0.001 | 1.30 (1.13–1.50) | <0.001 | 1.18 (1.01–1.38) | 0.039 |

First models adjust for completed assessments; the genetic model also adjusts for sex and ten ancestry principal components. Sociodemographic models add sex, ethnicity, maternal education and social class; fully adjusted models mutually include non-genetic exposures. Maternal education reference: A level or above. Count exposures are per additional occasion, trauma or SD; dashes indicate estimates not available. IRR, incidence rate ratio; RRR, relative risk ratio; CI, confidence interval; EA, educational attainment; KS2, Key Stage 2; SD, standard deviation; SDQ-C, Strengths and Difficulties Questionnaire conduct-problems subscale; SDQ-H, Strengths and Difficulties Questionnaire hyperactivity subscale; SMFQ, Short Mood and Feelings Questionnaire.

### Supplementary Table S4

**Supplementary Table 4.** Associations with age at first observed NEET, complete-case analysis (n=4,688).

| Exposure | Unadjusted | P | Sociodemographic | P | Fully adjusted | P |
|---|---|---|---|---|---|---|
| ***First observed NEET at 16–20 years*** | | | | | | |
| EA polygenic score (per SD) | 0.85 (0.77–0.93) | 0.001 | — | — | — | — |
| Female sex | 0.83 (0.67–1.02) | 0.079 | 0.81 (0.65–0.99) | 0.045 | 0.76 (0.61–0.95) | 0.015 |
| Non-White ethnicity | 1.36 (0.82–2.27) | 0.230 | 1.42 (0.85–2.36) | 0.180 | 1.34 (0.79–2.26) | 0.274 |
| Maternal education: O level | 1.35 (1.07–1.69) | 0.011 | 1.31 (1.01–1.70) | 0.041 | 1.24 (0.95–1.62) | 0.107 |
| Maternal education: below O level | 1.60 (1.22–2.11) | <0.001 | 1.57 (1.16–2.14) | 0.004 | 1.39 (1.01–1.92) | 0.043 |
| Lower maternal social class | 1.28 (1.04–1.57) | 0.022 | 1.08 (0.85–1.38) | 0.524 | 1.04 (0.81–1.33) | 0.784 |
| Maternal-depression occasions | 1.03 (0.96–1.10) | 0.462 | 1.02 (0.95–1.10) | 0.575 | 0.90 (0.83–0.98) | 0.015 |
| Childhood financial-problem occasions | 1.15 (1.08–1.23) | <0.001 | 1.14 (1.06–1.22) | <0.001 | 1.09 (1.01–1.17) | 0.023 |
| Childhood physical illness | 1.05 (0.71–1.56) | 0.798 | 1.02 (0.69–1.51) | 0.932 | 0.84 (0.56–1.26) | 0.407 |
| Childhood traumas | 1.38 (1.27–1.50) | <0.001 | 1.38 (1.26–1.50) | <0.001 | 1.26 (1.15–1.39) | <0.001 |
| KS2 attainment (per SD) | 0.77 (0.69–0.86) | <0.001 | 0.80 (0.71–0.90) | <0.001 | 0.88 (0.78–1.00) | 0.049 |
| Childhood conduct-problem occasions (SDQ-C) | 1.42 (1.27–1.59) | <0.001 | 1.42 (1.27–1.59) | <0.001 | 1.21 (1.06–1.38) | 0.006 |
| Childhood hyperactivity-problem occasions (SDQ-H) | 1.26 (1.16–1.38) | <0.001 | 1.25 (1.14–1.36) | <0.001 | 1.07 (0.96–1.19) | 0.239 |
| Adolescent-depression occasions (SMFQ) | 1.54 (1.35–1.76) | <0.001 | 1.57 (1.38–1.80) | <0.001 | 1.36 (1.18–1.57) | <0.001 |
| ***First observed NEET at 21–24 years*** | | | | | | |
| EA polygenic score (per SD) | 1.05 (0.94–1.16) | 0.387 | — | — | — | — |
| Female sex | 0.84 (0.67–1.06) | 0.146 | 0.86 (0.68–1.08) | 0.194 | 0.86 (0.68–1.10) | 0.229 |
| Non-White ethnicity | 1.44 (0.82–2.51) | 0.202 | 1.41 (0.81–2.46) | 0.229 | 1.41 (0.80–2.47) | 0.231 |
| Maternal education: O level | 0.87 (0.68–1.11) | 0.270 | 0.96 (0.73–1.27) | 0.790 | 0.97 (0.73–1.28) | 0.813 |
| Maternal education: below O level | 0.71 (0.50–1.02) | 0.061 | 0.80 (0.55–1.18) | 0.263 | 0.81 (0.55–1.21) | 0.303 |
| Lower maternal social class | 0.79 (0.63–0.99) | 0.041 | 0.84 (0.65–1.10) | 0.202 | 0.84 (0.64–1.09) | 0.190 |
| Maternal-depression occasions | 1.04 (0.96–1.12) | 0.348 | 1.04 (0.96–1.13) | 0.296 | 1.01 (0.93–1.10) | 0.814 |
| Childhood financial-problem occasions | 0.99 (0.91–1.08) | 0.839 | 1.01 (0.93–1.10) | 0.831 | 0.98 (0.90–1.07) | 0.682 |
| Childhood physical illness | 1.24 (0.82–1.88) | 0.316 | 1.23 (0.81–1.88) | 0.326 | 1.16 (0.76–1.78) | 0.491 |
| Childhood traumas | 1.12 (1.01–1.23) | 0.026 | 1.12 (1.02–1.24) | 0.022 | 1.07 (0.96–1.19) | 0.210 |
| KS2 attainment (per SD) | 1.03 (0.89–1.18) | 0.731 | 0.98 (0.84–1.13) | 0.736 | 1.03 (0.89–1.20) | 0.673 |
| Childhood conduct-problem occasions (SDQ-C) | 1.15 (0.99–1.34) | 0.073 | 1.16 (0.99–1.35) | 0.062 | 1.04 (0.87–1.24) | 0.659 |
| Childhood hyperactivity-problem occasions (SDQ-H) | 1.15 (1.03–1.28) | 0.011 | 1.15 (1.03–1.28) | 0.012 | 1.11 (0.98–1.25) | 0.106 |
| Adolescent-depression occasions (SMFQ) | 1.23 (1.05–1.45) | 0.011 | 1.26 (1.07–1.49) | 0.005 | 1.21 (1.02–1.43) | 0.031 |
| ***First observed NEET at 25–33 years*** | | | | | | |
| EA polygenic score (per SD) | 0.84 (0.72–0.97) | 0.022 | — | — | — | — |
| Female sex | 2.06 (1.42–2.99) | <0.001 | 2.07 (1.43–3.00) | <0.001 | 2.10 (1.43–3.08) | <0.001 |
| Non-White ethnicity | 1.41 (0.65–3.08) | 0.388 | 1.43 (0.65–3.14) | 0.372 | 1.32 (0.59–2.91) | 0.500 |
| Maternal education: O level | 1.10 (0.78–1.56) | 0.575 | 1.07 (0.72–1.59) | 0.733 | 1.02 (0.68–1.52) | 0.924 |
| Maternal education: below O level | 0.96 (0.60–1.54) | 0.863 | 0.90 (0.53–1.50) | 0.674 | 0.77 (0.45–1.32) | 0.343 |
| Lower maternal social class | 1.05 (0.76–1.45) | 0.757 | 1.02 (0.70–1.49) | 0.903 | 0.97 (0.66–1.42) | 0.870 |
| Maternal-depression occasions | 1.17 (1.07–1.28) | <0.001 | 1.17 (1.06–1.28) | <0.001 | 1.11 (1.00–1.23) | 0.045 |
| Childhood financial-problem occasions | 1.10 (0.99–1.23) | 0.082 | 1.09 (0.98–1.22) | 0.129 | 1.01 (0.90–1.14) | 0.873 |
| Childhood physical illness | 1.04 (0.55–1.94) | 0.909 | 1.13 (0.60–2.13) | 0.702 | 0.94 (0.49–1.78) | 0.849 |
| Childhood traumas | 1.30 (1.14–1.48) | <0.001 | 1.30 (1.14–1.49) | <0.001 | 1.23 (1.07–1.42) | 0.005 |
| KS2 attainment (per SD) | 0.77 (0.64–0.92) | 0.004 | 0.76 (0.63–0.92) | 0.005 | 0.82 (0.67–0.99) | 0.043 |
| Childhood conduct-problem occasions (SDQ-C) | 0.99 (0.77–1.27) | 0.954 | 0.99 (0.77–1.28) | 0.956 | 0.77 (0.58–1.02) | 0.070 |
| Childhood hyperactivity-problem occasions (SDQ-H) | 1.16 (1.00–1.34) | 0.051 | 1.21 (1.04–1.40) | 0.012 | 1.15 (0.97–1.36) | 0.106 |
| Adolescent-depression occasions (SMFQ) | 1.36 (1.10–1.69) | 0.004 | 1.29 (1.04–1.60) | 0.021 | 1.15 (0.92–1.45) | 0.216 |

First models adjust for completed assessments; the genetic model also adjusts for sex and ten ancestry principal components. Sociodemographic models add sex, ethnicity, maternal education and social class; fully adjusted models mutually include non-genetic exposures. Maternal education reference: A level or above. Count exposures are per additional occasion, trauma or SD; dashes indicate estimates not available. IRR, incidence rate ratio; RRR, relative risk ratio; CI, confidence interval; EA, educational attainment; KS2, Key Stage 2; SD, standard deviation; SDQ-C, Strengths and Difficulties Questionnaire conduct-problems subscale; SDQ-H, Strengths and Difficulties Questionnaire hyperactivity subscale; SMFQ, Short Mood and Feelings Questionnaire.

### Supplementary Table S5

**Supplementary Table 5.** Associations with pattern of observed NEET, relative to never-observed NEET. Imputed N=8130. Estimates are RRRs (95% CIs).

| Exposure | Unadjusted | P | Sociodemographic | P | Fully adjusted | P |
|---|---|---|---|---|---|---|
| ***Single Occasion NEET*** | | | | | | |
| EA polygenic score (per SD) | 0.95 (0.88–1.03) | 0.201 | — | — | — | — |
| Female sex | 0.94 (0.82–1.07) | 0.338 | 0.93 (0.82–1.06) | 0.267 | 0.90 (0.78–1.03) | 0.116 |
| Non-White ethnicity | 1.42 (1.07–1.89) | 0.016 | 1.42 (1.07–1.89) | 0.015 | 1.35 (1.02–1.80) | 0.037 |
| Maternal education: O level | 1.05 (0.91–1.22) | 0.504 | 1.09 (0.92–1.29) | 0.336 | 1.04 (0.87–1.24) | 0.679 |
| Maternal education: below O level | 1.14 (0.96–1.36) | 0.146 | 1.18 (0.96–1.44) | 0.111 | 1.05 (0.85–1.30) | 0.630 |
| Lower maternal social class | 1.01 (0.88–1.16) | 0.873 | 0.96 (0.81–1.13) | 0.596 | 0.92 (0.78–1.09) | 0.341 |
| Maternal-depression occasions | 1.07 (1.02–1.11) | 0.002 | 1.06 (1.02–1.11) | 0.003 | 1.00 (0.96–1.05) | 0.833 |
| Childhood financial-problem occasions | 1.09 (1.04–1.14) | <0.001 | 1.08 (1.04–1.13) | <0.001 | 1.04 (1.00–1.09) | 0.071 |
| Childhood physical illness | 0.98 (0.75–1.29) | 0.903 | 0.97 (0.74–1.27) | 0.843 | 0.88 (0.67–1.15) | 0.346 |
| Childhood traumas | 1.18 (1.12–1.25) | <0.001 | 1.18 (1.11–1.24) | <0.001 | 1.10 (1.03–1.17) | 0.003 |
| KS2 attainment (per SD) | 0.85 (0.79–0.92) | <0.001 | 0.85 (0.78–0.92) | <0.001 | 0.90 (0.83–0.98) | 0.015 |
| Childhood conduct-problem occasions (SDQ-C) | 1.19 (1.11–1.28) | <0.001 | 1.19 (1.11–1.28) | <0.001 | 1.07 (0.98–1.16) | 0.146 |
| Childhood hyperactivity-problem occasions (SDQ-H) | 1.15 (1.09–1.21) | <0.001 | 1.14 (1.08–1.21) | <0.001 | 1.05 (0.99–1.13) | 0.110 |
| Adolescent-depression occasions (SMFQ) | 1.24 (1.14–1.35) | <0.001 | 1.26 (1.15–1.37) | <0.001 | 1.17 (1.06–1.28) | 0.001 |
| ***Sustained NEET*** | | | | | | |
| EA polygenic score (per SD) | 0.87 (0.75–1.01) | 0.059 | — | — | — | — |
| Female sex | 1.17 (0.92–1.50) | 0.196 | 1.11 (0.87–1.42) | 0.399 | 1.05 (0.80–1.37) | 0.736 |
| Non-White ethnicity | 1.20 (0.69–2.08) | 0.519 | 1.17 (0.68–2.04) | 0.569 | 1.06 (0.60–1.88) | 0.839 |
| Maternal education: O level | 1.14 (0.85–1.54) | 0.373 | 1.18 (0.84–1.66) | 0.331 | 1.01 (0.71–1.43) | 0.959 |
| Maternal education: below O level | 2.25 (1.68–3.00) | <0.001 | 2.31 (1.64–3.27) | <0.001 | 1.54 (1.06–2.24) | 0.023 |
| Lower maternal social class | 1.21 (0.93–1.58) | 0.153 | 0.93 (0.67–1.28) | 0.659 | 0.80 (0.57–1.12) | 0.195 |
| Maternal-depression occasions | 1.18 (1.11–1.26) | <0.001 | 1.16 (1.09–1.24) | <0.001 | 0.99 (0.92–1.07) | 0.864 |
| Childhood financial-problem occasions | 1.22 (1.13–1.31) | <0.001 | 1.18 (1.10–1.28) | <0.001 | 1.04 (0.96–1.13) | 0.334 |
| Childhood physical illness | 1.83 (1.23–2.72) | 0.003 | 1.85 (1.24–2.76) | 0.003 | 1.30 (0.85–1.98) | 0.223 |
| Childhood traumas | 1.61 (1.47–1.76) | <0.001 | 1.60 (1.46–1.75) | <0.001 | 1.31 (1.18–1.45) | <0.001 |
| KS2 attainment (per SD) | 0.51 (0.46–0.58) | <0.001 | 0.53 (0.47–0.61) | <0.001 | 0.65 (0.56–0.75) | <0.001 |
| Childhood conduct-problem occasions (SDQ-C) | 1.69 (1.54–1.86) | <0.001 | 1.67 (1.52–1.83) | <0.001 | 1.27 (1.12–1.45) | <0.001 |
| Childhood hyperactivity-problem occasions (SDQ-H) | 1.46 (1.35–1.58) | <0.001 | 1.47 (1.35–1.59) | <0.001 | 1.10 (0.98–1.23) | 0.102 |
| Adolescent-depression occasions (SMFQ) | 1.77 (1.56–2.00) | <0.001 | 1.76 (1.55–2.01) | <0.001 | 1.38 (1.20–1.60) | <0.001 |
| ***Recurrent NEET*** | | | | | | |
| EA polygenic score (per SD) | 0.79 (0.64–0.96) | 0.018 | — | — | — | — |
| Female sex | 1.53 (1.09–2.16) | 0.015 | 1.46 (1.03–2.06) | 0.033 | 1.34 (0.93–1.92) | 0.118 |
| Non-White ethnicity | 2.28 (1.29–4.02) | 0.005 | 2.25 (1.27–3.98) | 0.005 | 2.08 (1.17–3.72) | 0.013 |
| Maternal education: O level | 1.18 (0.81–1.72) | 0.395 | 1.16 (0.75–1.79) | 0.502 | 1.05 (0.68–1.62) | 0.833 |
| Maternal education: below O level | 2.01 (1.35–3.00) | <0.001 | 1.90 (1.19–3.03) | 0.007 | 1.54 (0.94–2.50) | 0.084 |
| Lower maternal social class | 1.28 (0.91–1.78) | 0.153 | 1.04 (0.69–1.55) | 0.863 | 0.96 (0.64–1.45) | 0.862 |
| Maternal-depression occasions | 1.06 (0.96–1.18) | 0.234 | 1.04 (0.94–1.15) | 0.453 | 0.93 (0.83–1.04) | 0.197 |
| Childhood financial-problem occasions | 1.17 (1.06–1.29) | 0.002 | 1.13 (1.02–1.25) | 0.020 | 1.07 (0.96–1.19) | 0.247 |
| Childhood physical illness | 1.96 (1.19–3.24) | 0.008 | 2.05 (1.24–3.39) | 0.005 | 1.73 (1.03–2.89) | 0.039 |
| Childhood traumas | 1.40 (1.23–1.58) | <0.001 | 1.38 (1.22–1.56) | <0.001 | 1.23 (1.06–1.42) | 0.005 |
| KS2 attainment (per SD) | 0.68 (0.57–0.80) | <0.001 | 0.72 (0.60–0.86) | <0.001 | 0.80 (0.66–0.97) | 0.021 |
| Childhood conduct-problem occasions (SDQ-C) | 1.45 (1.25–1.67) | <0.001 | 1.42 (1.22–1.64) | <0.001 | 1.28 (1.06–1.54) | 0.010 |
| Childhood hyperactivity-problem occasions (SDQ-H) | 1.17 (1.02–1.34) | 0.023 | 1.18 (1.02–1.35) | 0.021 | 0.94 (0.79–1.12) | 0.499 |
| Adolescent-depression occasions (SMFQ) | 1.58 (1.33–1.87) | <0.001 | 1.53 (1.29–1.83) | <0.001 | 1.34 (1.11–1.63) | 0.003 |

First models adjust for completed assessments; the genetic model also adjusts for sex and ten ancestry principal components. Sociodemographic models add sex, ethnicity, maternal education and social class; fully adjusted models mutually include non-genetic exposures. Maternal education reference: A level or above. Count exposures are per additional occasion, trauma or SD; dashes indicate estimates not available. IRR, incidence rate ratio; RRR, relative risk ratio; CI, confidence interval; EA, educational attainment; KS2, Key Stage 2; SD, standard deviation; SDQ-C, Strengths and Difficulties Questionnaire conduct-problems subscale; SDQ-H, Strengths and Difficulties Questionnaire hyperactivity subscale; SMFQ, Short Mood and Feelings Questionnaire.

### Supplementary Table S6

**Supplementary Table 6.** Associations with pattern of observed NEET, complete-case analysis (n=4,688).

| Exposure | Unadjusted | P | Sociodemographic | P | Fully adjusted | P |
|---|---|---|---|---|---|---|
| ***Single Occasion NEET*** | | | | | | |
| EA polygenic score (per SD) | 0.95 (0.88–1.03) | 0.201 | — | — | — | — |
| Female sex | 0.86 (0.73–1.03) | 0.097 | 0.87 (0.73–1.03) | 0.106 | 0.86 (0.72–1.02) | 0.088 |
| Non-White ethnicity | 1.30 (0.85–2.00) | 0.224 | 1.31 (0.85–2.02) | 0.217 | 1.27 (0.82–1.96) | 0.281 |
| Maternal education: O level | 1.09 (0.91–1.31) | 0.356 | 1.11 (0.90–1.36) | 0.328 | 1.09 (0.88–1.34) | 0.425 |
| Maternal education: below O level | 0.88 (0.69–1.14) | 0.336 | 0.91 (0.69–1.19) | 0.483 | 0.88 (0.66–1.16) | 0.359 |
| Lower maternal social class | 0.99 (0.84–1.18) | 0.951 | 0.99 (0.81–1.20) | 0.881 | 0.96 (0.79–1.18) | 0.715 |
| Maternal-depression occasions | 1.03 (0.98–1.10) | 0.264 | 1.03 (0.98–1.10) | 0.258 | 0.98 (0.92–1.04) | 0.524 |
| Childhood financial-problem occasions | 1.07 (1.01–1.14) | 0.017 | 1.08 (1.02–1.15) | 0.012 | 1.05 (0.99–1.12) | 0.116 |
| Childhood physical illness | 0.94 (0.67–1.32) | 0.731 | 0.92 (0.66–1.30) | 0.642 | 0.85 (0.60–1.20) | 0.344 |
| Childhood traumas | 1.18 (1.10–1.27) | <0.001 | 1.18 (1.10–1.27) | <0.001 | 1.13 (1.05–1.23) | 0.002 |
| KS2 attainment (per SD) | 0.95 (0.86–1.06) | 0.375 | 0.94 (0.85–1.05) | 0.254 | 0.99 (0.89–1.11) | 0.883 |
| Childhood conduct-problem occasions (SDQ-C) | 1.12 (1.00–1.26) | 0.058 | 1.12 (1.00–1.26) | 0.053 | 0.99 (0.87–1.13) | 0.856 |
| Childhood hyperactivity-problem occasions (SDQ-H) | 1.15 (1.06–1.24) | <0.001 | 1.14 (1.05–1.24) | 0.001 | 1.10 (1.00–1.20) | 0.046 |
| Adolescent-depression occasions (SMFQ) | 1.28 (1.13–1.45) | <0.001 | 1.30 (1.15–1.47) | <0.001 | 1.22 (1.07–1.38) | 0.002 |
| ***Sustained NEET*** | | | | | | |
| EA polygenic score (per SD) | 0.87 (0.75–1.01) | 0.059 | — | — | — | — |
| Female sex | 1.26 (0.90–1.77) | 0.178 | 1.22 (0.87–1.72) | 0.246 | 1.18 (0.82–1.68) | 0.372 |
| Non-White ethnicity | 1.19 (0.52–2.76) | 0.680 | 1.22 (0.53–2.83) | 0.645 | 1.15 (0.48–2.74) | 0.751 |
| Maternal education: O level | 1.12 (0.78–1.61) | 0.547 | 1.27 (0.84–1.93) | 0.257 | 1.14 (0.75–1.75) | 0.534 |
| Maternal education: below O level | 1.96 (1.31–2.93) | 0.001 | 2.24 (1.41–3.54) | <0.001 | 1.70 (1.04–2.75) | 0.033 |
| Lower maternal social class | 1.00 (0.73–1.38) | 0.985 | 0.77 (0.53–1.13) | 0.180 | 0.70 (0.47–1.03) | 0.069 |
| Maternal-depression occasions | 1.17 (1.07–1.28) | <0.001 | 1.16 (1.06–1.27) | 0.001 | 1.04 (0.94–1.15) | 0.483 |
| Childhood financial-problem occasions | 1.12 (1.01–1.25) | 0.032 | 1.10 (0.99–1.23) | 0.088 | 0.98 (0.87–1.11) | 0.770 |
| Childhood physical illness | 1.24 (0.69–2.22) | 0.477 | 1.29 (0.72–2.33) | 0.390 | 0.93 (0.51–1.71) | 0.816 |
| Childhood traumas | 1.51 (1.34–1.71) | <0.001 | 1.50 (1.33–1.70) | <0.001 | 1.27 (1.10–1.46) | <0.001 |
| KS2 attainment (per SD) | 0.56 (0.48–0.66) | <0.001 | 0.58 (0.49–0.69) | <0.001 | 0.66 (0.56–0.80) | <0.001 |
| Childhood conduct-problem occasions (SDQ-C) | 1.64 (1.42–1.90) | <0.001 | 1.64 (1.42–1.90) | <0.001 | 1.27 (1.06–1.53) | 0.010 |
| Childhood hyperactivity-problem occasions (SDQ-H) | 1.42 (1.26–1.60) | <0.001 | 1.45 (1.28–1.63) | <0.001 | 1.13 (0.97–1.31) | 0.126 |
| Adolescent-depression occasions (SMFQ) | 1.73 (1.44–2.08) | <0.001 | 1.71 (1.42–2.06) | <0.001 | 1.40 (1.14–1.71) | 0.001 |
| ***Recurrent NEET*** | | | | | | |
| EA polygenic score (per SD) | 0.79 (0.64–0.96) | 0.018 | — | — | — | — |
| Female sex | 1.34 (0.86–2.09) | 0.189 | 1.31 (0.84–2.03) | 0.235 | 1.29 (0.81–2.04) | 0.279 |
| Non-White ethnicity | 2.45 (1.11–5.44) | 0.027 | 2.51 (1.13–5.58) | 0.024 | 2.45 (1.08–5.52) | 0.031 |
| Maternal education: O level | 1.11 (0.70–1.75) | 0.651 | 0.98 (0.58–1.64) | 0.925 | 0.95 (0.56–1.60) | 0.838 |
| Maternal education: below O level | 1.60 (0.94–2.73) | 0.086 | 1.36 (0.75–2.47) | 0.319 | 1.31 (0.70–2.44) | 0.393 |
| Lower maternal social class | 1.40 (0.92–2.12) | 0.115 | 1.33 (0.81–2.17) | 0.257 | 1.28 (0.78–2.11) | 0.329 |
| Maternal-depression occasions | 1.02 (0.88–1.18) | 0.831 | 1.00 (0.86–1.16) | 0.988 | 0.89 (0.76–1.04) | 0.154 |
| Childhood financial-problem occasions | 1.13 (0.98–1.29) | 0.083 | 1.09 (0.95–1.25) | 0.234 | 1.04 (0.89–1.21) | 0.621 |
| Childhood physical illness | 2.26 (1.24–4.14) | 0.008 | 2.38 (1.30–4.38) | 0.005 | 2.09 (1.12–3.88) | 0.020 |
| Childhood traumas | 1.49 (1.27–1.75) | <0.001 | 1.47 (1.26–1.73) | <0.001 | 1.38 (1.15–1.64) | <0.001 |
| KS2 attainment (per SD) | 0.85 (0.67–1.07) | 0.169 | 0.89 (0.69–1.15) | 0.372 | 1.01 (0.78–1.31) | 0.940 |
| Childhood conduct-problem occasions (SDQ-C) | 1.45 (1.18–1.79) | <0.001 | 1.45 (1.17–1.79) | <0.001 | 1.26 (0.97–1.62) | 0.080 |
| Childhood hyperactivity-problem occasions (SDQ-H) | 1.21 (1.01–1.44) | 0.043 | 1.22 (1.01–1.46) | 0.036 | 1.01 (0.81–1.26) | 0.933 |
| Adolescent-depression occasions (SMFQ) | 1.62 (1.27–2.06) | <0.001 | 1.58 (1.24–2.01) | <0.001 | 1.34 (1.03–1.74) | 0.028 |

First models adjust for completed assessments; the genetic model also adjusts for sex and ten ancestry principal components. Sociodemographic models add sex, ethnicity, maternal education and social class; fully adjusted models mutually include non-genetic exposures. Maternal education reference: A level or above. Count exposures are per additional occasion, trauma or SD; dashes indicate estimates not available. IRR, incidence rate ratio; RRR, relative risk ratio; CI, confidence interval; EA, educational attainment; KS2, Key Stage 2; SD, standard deviation; SDQ-C, Strengths and Difficulties Questionnaire conduct-problems subscale; SDQ-H, Strengths and Difficulties Questionnaire hyperactivity subscale; SMFQ, Short Mood and Feelings Questionnaire.

### Supplementary Table S7

Supplementary Table S7. Measurement of childhood exposure variables: age(s) assessed and instrument or source for each exposure used in the main analyses.

| Exposure | Age(s)/timing assessed | Instrument / source |
|---|---|---|
| Sex assigned at birth | Birth | Birth record |
| Ethnicity | Birth | Parent-reported |
| Maternal education and social class | Birth | Parent-reported |
| Maternal-depression count | 8 occasions, 32 weeks' gestation to 8 years | Edinburgh Postnatal Depression Scale (EPDS) |
| Childhood financial-problems count | 5 occasions, 32 weeks' gestation to 7 years | Parent-reported questionnaire |
| Childhood physical illness | Up to age 16 | Parent-reported questionnaire |
| Childhood trauma count | Up to age 17 | Retrospective questionnaire |
| Key Stage 2 (KS2) attainment | Age 10–11 | Standardised routine UK school assessment |
| Childhood conduct-problems count | 5 occasions, ages 6.5–16 years | Strengths and Difficulties Questionnaire (SDQ), conduct-problems subscale |
| Childhood hyperactivity count | 5 occasions, ages 6.5–16 years | Strengths and Difficulties Questionnaire (SDQ), hyperactivity subscale |
| Adolescent-depression occasions | 4 occasions, ages 10–16 years | Short Mood and Feelings Questionnaire (SMFQ) |
| Educational-attainment polygenic score | Derived from genotype (not age-specific) | Derived from a genome-wide association study of educational attainment |
